# High vaccine effectiveness against severe Covid-19 in the elderly in Finland before and after the emergence of Omicron

**DOI:** 10.1101/2022.03.11.22272140

**Authors:** Ulrike Baum, Eero Poukka, Tuija Leino, Terhi Kilpi, Hanna Nohynek, Arto A. Palmu

## Abstract

**Background:** The elderly are highly vulnerable to severe Covid-19. Waning immunity and emergence of Omicron have caused concerns about reduced effectiveness of Covid-19 vaccines. The objective was to estimate vaccine effectiveness (VE) against severe Covid-19 among the elderly.

**Methods:** This nationwide, register-based cohort analysis included all residents aged 70 years and over in Finland. The follow-up started on December 27, 2020, and ended on March 31, 2022. The outcomes of interest were Covid-19-related hospitalization and intensive care unit (ICU) admission timely associated with SARS-CoV-2 infection. VE was estimated as one minus the hazard ratio comparing the vaccinated and unvaccinated and taking into account time since vaccination. Omicron-specific VE was evaluated as the effectiveness observed since January 1, 2022.

**Results:** The cohort included 896220 individuals. Comirnaty (BioNTech/Pfizer) VE against Covid-19-related hospitalization was 93% (95% CI 89%–95%) and 85% (95% CI 82%–87%) 14–90 and 91–180 days after the second dose; VE increased to 95% (95% CI 94%–96%) 14–60 days after the third dose. VE of other homologous and heterologous three dose series was similar. Protection against severe Covid-19 requiring ICU treatment was even better. Since January 1, 2022, Comirnaty VE was 98% (95% CI 92%–99%) and 92% (95% CI 87%–95%) 14–90 and 91–180 days after the second and 98% (95% CI 95%–99%) 14–60 days after the third dose.

**Conclusions:** VE against severe Covid-19 is high among the elderly. It waned slightly after two doses, but a third restored the protection. VE against severe Covid-19 remained high even after the emergence of Omicron.

## Background

The elderly are highly vulnerable to severe coronavirus disease 2019 (Covid-19) (1) and, therefore, protecting them is essential to reduce the disease burden caused by the severe acute respiratory syndrome coronavirus 2 (SARS-CoV-2). Covid-19 vaccinations prevent SARS-CoV-2 infections and decrease the number of Covid-19 hospitalizations in a population (2–7).

As many other European countries, Finland has mainly used two mRNA Covid-19 vaccines, Comirnaty and Spikevax, and one adenovirus vector Covid-19 vaccine, Vaxzevria (Table 1). In September 2021, Finland started its booster vaccination campaign to improve the protection especially for the elderly (8). The initial recommendation was to adhere to homologous vaccine series, but lately with increasing evidence of good effectiveness (9,10), heterologous series have also been encouraged.

**Table 1.** Covid-19 vaccines under study

| Trade name | International nonproprietary name | Trial name | Manufacturer | Vaccine type | Available in Finland |
| --- | --- | --- | --- | --- | --- |
| Comirnaty | Tozinameran | BNT162b2 | BioNTech/Pfizer | mRNA vaccine | Since December 27, 2020 |
| Spikevax | Elasomeran | mRNA-1273 | Moderna | mRNA vaccine | Since January 20, 2021 |
| Vaxzevria | ChAdOx1-S [recombinant] | AZD1222 | Oxford/AstraZeneca | Adenovirus vector vaccine | From February 10 to November 30, 2021 |

Vaccine effectiveness (VE) generally describes the protective direct effect of vaccination. In the elderly, VE against severe Covid-19 is high after the second dose, but it decreases during the following six months (4,11–14). A third dose increased the VE against infection among adults in multiple observational studies (11,15–18). However, only a few studies have investigated how well the third dose boosts the VE against severe disease among the elderly (17,19,20). Furthermore, VE may decrease because of the recently emerged Omicron variant, which is more capable of evading both natural and vaccine-induced immunity than previous variants (15,16,21–23). Knowledge of VE against severe Covid-19 caused by Omicron is still limited (16,21,23–25) and discussion of the need for further booster doses in the elderly has commenced.

The objective of this study was to estimate the effectiveness of Covid-19 vaccines against severe Covid-19 requiring hospitalization or intensive care unit (ICU) treatment in the elderly in Finland after two and three doses. The effects of time since vaccination, different vaccine series, age and presence of comorbidities on VE were of particular interest. In addition, this study aimed to evaluate the impact of the recently emerged Omicron variant on VE.

## Methods

To estimate the effectiveness of Covid-19 vaccines in the elderly population in Finland, we conducted a nationwide, register-based cohort analysis starting on December 27, 2020, and ending on March 31, 2022. The study population was defined as all individuals aged 70 years and over at the beginning of the study and registered in the Population Information System as resident in Finland since January 1, 2020. The unique personal identity code assigned to all permanent residents in Finland allowed linking individual-level data from different sources.

The primary outcome was Covid-19-related hospital admission timely associated with a laboratory-confirmed (by polymerase chain reaction or antigen detection assay) SARS-CoV-2 infection. Using the Care Register for Health Care, which records data on all patients discharged from inpatient care in Finland, we defined Covid-19-related hospitalization as any inpatient encounter with a primary diagnosis of Covid-19 (International Classification of Diseases, 10^th^ revision: U07.1, U07.2), acute respiratory tract infection (J00– J22, J46) or severe complication of lower respiratory tract infections (J80–84, J85.1, J86). A hospitalization was considered timely associated with an infection recorded in the National Infectious Diseases Register if the positive specimen was collected up to 14 days before or seven days after the hospital admission.

The secondary outcome was Covid-19-related ICU admission. ICU admissions were identified from the Finnish Intensive Care Consortium’s Quality Register for Intensive Care, which records data on all patients treated in an ICU in Finland. We considered any admission as Covid-19 related if it was marked by the treating physician as due to Covid-19 and if the patient was laboratory-confirmed SARS-CoV-2 positive during the stay.

The exposure was Covid-19 vaccination recorded in the National Vaccination Register, which covers the whole population irrespective of whether they are served by public or private primary health care providers. We distinguished between the three vaccine brands Comirnaty, Spikevax and Vaxzevria and the number of administered doses. The time since vaccination was taken into account by categorizing the time since the last dose using the following cut points: days 21 and 84 after the first dose, days 14, 91 and 181 after the second dose, and days 14 and 61 after the third dose. Thus, a vaccinee’s exposure state changed over time. Being unvaccinated was the reference state.

We considered age, sex, region of residence, residence in a long-term care facility, influenza vaccination in 2019–2020, number of nights hospitalized between 2015 and 2019 and presence of predisposing comorbidities or medical therapies as confounders. The first three confounders were taken from the Population Information System. Information on whether a subject was in long-term care at the beginning of the study or vaccinated against influenza in the last pre-pandemic season were collected from the Care Register for Social Care and the National Vaccination Register, respectively. We used the data in the Care Register for Health Care from 2015 onwards to count the number of nights hospitalized between 2015 and 2019 and to assess the presence of comorbidities or medical therapies that predispose to severe Covid-19 according to a recent study of predictors of Covid-19 hospitalization (26). We completed the collection of data on predisposing comorbidities and medical therapies with primary health care records and prescription data as outlined in Supplementary Tables 1-2.

Each study subject was considered at risk of the primary and secondary outcomes from the beginning of the study until the first occurrence of any of the following events: outcome of interest, death, day 14 after any laboratory-confirmed SARS-CoV-2 infection, vaccination with an unidentified vaccine, a heterologous second vaccination, vaccination with the third dose prior to the start of the booster campaign (approximated by September 17, 2021), a fourth vaccination, or end of study. All those events other than the outcome of interest led to censoring before or at the end of the study.

Using Cox regression with time in the study as the underlying time scale, we compared the hazard of the two outcomes in vaccinated study subjects with the corresponding hazard in the unvaccinated. The effect measure of interest was VE, quantified as one minus the hazard ratio adjusted for the seven confounders categorized as outlined in Table 2. For each VE estimate, we computed either the 95% Wald confidence interval (CI) or, if there were no cases in one of the two groups, the p-value of the likelihood-ratio test. We stratified the analysis by age group and presence of comorbidities or medical therapies. Because of the emergence of Omicron (Supplementary Figure 1), we also estimated VE by calendar time, conducting a separate regression analysis for each quarter.

**Table 2.** Distribution of person-years in the study of vaccine effectiveness against Covid-19 hospitalization.

|  | Not vaccinated, person-years (%) | Vaccinated, person-years (%) |
| --- | --- | --- |
| **Age in years** |  |  |
| 70-79 | 173 830 (24) | 535 938 (76) |
| 80-89 | 57 939 (19) | 246 105 (81) |
| 90-115 | 13 068 (22) | 47 634 (78) |
| **Sex** |  |  |
| Male | 105 628 (23) | 350 917 (77) |
| Female | 139 209 (23) | 478 761 (77) |
| **Region of residence** |  |  |
| Helsinki-Uusimaa | 56 435 (22) | 202 126 (78) |
| Åland | 1145 (19) | 4786 (81) |
| Northern and Eastern Finland | 61 478 (23) | 205 852 (77) |
| Southern Finland | 59 279 (23) | 197 393 (77) |
| Western Finland | 66 501 (23) | 219 521 (77) |
| **In long-term care** |  |  |
| No | 233 993 (23) | 779 369 (77) |
| Yes | 10 844 (18) | 50 309 (82) |
| **Influenza vaccination in 2019–2020** |  |  |
| No | 133 875 (29) | 335 681 (71) |
| Yes | 110 963 (18) | 493 997 (82) |
| **Nights hospitalized between 2015 and 2019** |  |  |
| 0 | 132 975 (24) | 429 964 (76) |
| 1-5 | 50 008 (22) | 181 812 (78) |
| 6-20 | 35 615 (22) | 126 826 (78) |
| 21+ | 26 240 (22) | 91 077 (78) |
| **Presence of comorbidities or medical therapies<sup>a</sup>** |  |  |
| No predisposing comorbidities | 84 412 (24) | 261 428 (76) |
| Only moderately predisposing comorbidities or medical therapies | 65 980 (22) | 232 391 (78) |
| At least one highly predisposing comorbidity or medical therapy | 94 445 (22) | 335 859 (78) |
| **Presence of particular comorbidities or medical therapies<sup>a</sup>** |  |  |
| Severe heart disease | 93 843 (22) | 337 889 (78) |
| Type 2 diabetes mellitus | 52 035 (22) | 182 964 (78) |
| Immunosuppressive medication | 35 443 (22) | 127 002 (78) |
| Actively treated cancer | 31 410 (22) | 114 354 (78) |
| Severe chronic respiratory disease | 26 198 (22) | 93 213 (78) |
| Neurological condition affecting breathing | 22 431 (22) | 79 939 (78) |
| Autoimmune disease | 17 062 (22) | 60 089 (78) |
| Type 1 diabetes mellitus or adrenal insufficiency | 13 627 (23) | 46 922 (77) |
| Sleep apnea | 11 867 (21) | 43 736 (79) |
| Severe kidney disease | 5548 (22) | 19 162 (78) |
| Psychotic disease | 3833 (29) | 9282 (71) |
| Severe chronic liver disease | 1165 (25) | 3568 (75) |
| Organ or stem cell transplantation | 560 (22) | 1962 (78) |
| Severe disorder of the immune system | 398 (23) | 1369 (77) |
| Down syndrome | 3 (19) | 13 (81) |

<sup>a</sup> See study of predictors of Covid-19 hospitalization (26) and Supplementary Tables 1-2.
This tabulation uses a binary vaccination status without distinguishing between vaccine brands, number of doses and time since vaccination.

To rule out residual confounding, we quantified the association between Covid-19 vaccination and a potential negative control outcome: inpatient encounters due to injury, poisoning and certain other consequences of external causes (International Classification of Diseases, 10^th^ revision: S00–T98) recorded in the Care Register for Health Care. Each study subject was considered at risk of this outcome from the beginning of the study until the first occurrence of any of the following events: injury, death, vaccination with an unidentified vaccine or a heterologous two-dose series, vaccination with the third dose prior to September 17, 2021, or with a fourth dose, or end of study. We compared the hazard of the control outcome in vaccinated study subjects with the corresponding hazard in the unvaccinated using Cox regression and expected to find no difference between the groups.

The significance level was set to 5%. The validity of the proportional hazards assumption was examined visually by plotting the cumulative hazards over time. All analyses were performed in R 4.1.2 (R Foundation for Statistical Computing, Vienna, Austria). Results concerning the first dose, less common (less than 2000 person-years) vaccine series and the first two weeks after vaccination are presented in the supplementary data only.

## Results

The study cohort included 896 220 individuals aged 70 years and over, and thus 99.8% of the elderly population in Finland (Supplementary Tables 3-4). At the end of the study, only 5% of those still considered at risk of Covid-19 hospitalization were unvaccinated; the majority (6% and 65%) were double- or triple-vaccinated with Comirnaty (Supplementary Table 5, Supplementary Figure 2). The median length of the dosing interval between the first and second dose and the second and third dose was 84 (interquartile range, 84–84) and 188 (interquartile range, 181–199) days, respectively. At the end of the study, the triple vaccinated who were still considered at risk of Covid-19 hospitalization had received their third dose, on average, 106 (interquartile range, 93–120) days ago. Table 2 shows the distribution of person-years by vaccination status. The younger and community-dwelling elderly contributed proportionally less vaccinated person-time than older individuals or those in long-term care. The younger and community-dwelling elderly had, however, received their first dose later than older individuals and those in long-term care (Supplementary Table 5).

We observed 2234 Covid-19 hospitalizations and 296 ICU admissions, of which 793 and 159 were among the unvaccinated (Table 3, Supplementary Figure 3, Supplementary Tables 6-7). In the first 14–90 days since the second dose of Comirnaty, the VE against hospitalization was 93% (95% CI 89%–95%). In the following 90 days, VE decreased to 85% (95% CI 82%–87%) and subsequently rose to 95% (95% CI 94%–96%) in the first 14–60 days since the third dose (Figure 1, Supplementary Table 8). The point estimates of VE against ICU admission were even higher (Figure 1, Supplementary Table 9). The only vaccine series with a statistically significantly lower VE than the corresponding homologous Comirnaty series was the two-dose homologous Vaxzevria series whose effectiveness against hospitalization was 74% (95% CI 60%–83%) in the first 91–180 days.

**Table 3.** Cases, person-years and cumulative risk (per 100 000) by vaccine, dose and days since last vaccination.

|  | Covid-19-related hospital admission |  |  | Covid-19-related ICU admission |  |  |
| --- | --- | --- | --- | --- | --- | --- |
|  | Cases | Person-years | Risk<sup>a</sup> | Cases | Person-years | Risk<sup>a</sup> |
| Not vaccinated | 793 | 244 838 | 1313 | 159 | 244 856 | 238 |
| Comirnaty + Comirnaty 14-90 | 33 | 144 412 | 108 | <5 | 144 413 | 8 |
| Comirnaty + Comirnaty 91-180 | 210 | 158 116 | 277 | 26 | 158 121 | 29 |
| Comirnaty + Comirnaty 181+ | 200 | 31 354 | 457 | 22 | 31 360 | 55 |
| Comirnaty + Comirnaty + Comirnaty 14-60 | 107 | 69 468 | 84 | 9 | 69 472 | 3 |
| Comirnaty + Comirnaty + Comirnaty 61+ | 359 | 74 262 | 125 | 26 | 74 271 | 55 |
| Comirnaty + Comirnaty + Spikevax 14-60 | 28 | 9222 | 118 | 5 | 9222 | 13 |
| Comirnaty + Comirnaty + Spikevax 61+ | 39 | 5601 | 325 | <5 | 5602 | 4 |
| Spikevax + Spikevax 14-90 | 5 | 16 589 | 201 | 0 | 16 589 | 0 |
| Spikevax + Spikevax 91-180 | 34 | 17 796 | 277 | <5 | 17 797 | 45 |
| Spikevax + Spikevax 181+ | 23 | 3315 | 433 | <5 | 3316 | 30 |
| Spikevax + Spikevax + Comirnaty 14-60 | 6 | 2167 | 137 | 0 | 2167 | 0 |
| Spikevax + Spikevax + Comirnaty 61+ | 10 | 2092 | 70 | 0 | 2092 | 0 |
| Spikevax + Spikevax + Spikevax 14-60 | 8 | 6630 | 27 | 0 | 6630 | 0 |
| Spikevax + Spikevax + Spikevax 61+ | 41 | 5948 | 109 | 0 | 5949 | 0 |
| Vaxzevria + Vaxzevria 14-90 | <5 | 8248 | 16 | <5 | 8248 | 10 |
| Vaxzevria + Vaxzevria 91-180 | 23 | 9067 | 205 | 5 | 9067 | 13 |
| Vaxzevria + Vaxzevria + Comirnaty 14-60 | <5 | 3044 | 24 | 0 | 3044 | 0 |
| Vaxzevria + Vaxzevria + Comirnaty 61+ | 12 | 2434 | 363 | <5 | 2434 | 307 |

<sup>a</sup> Estimated based on the Kaplan-Meier estimator.

**Figure 1.**
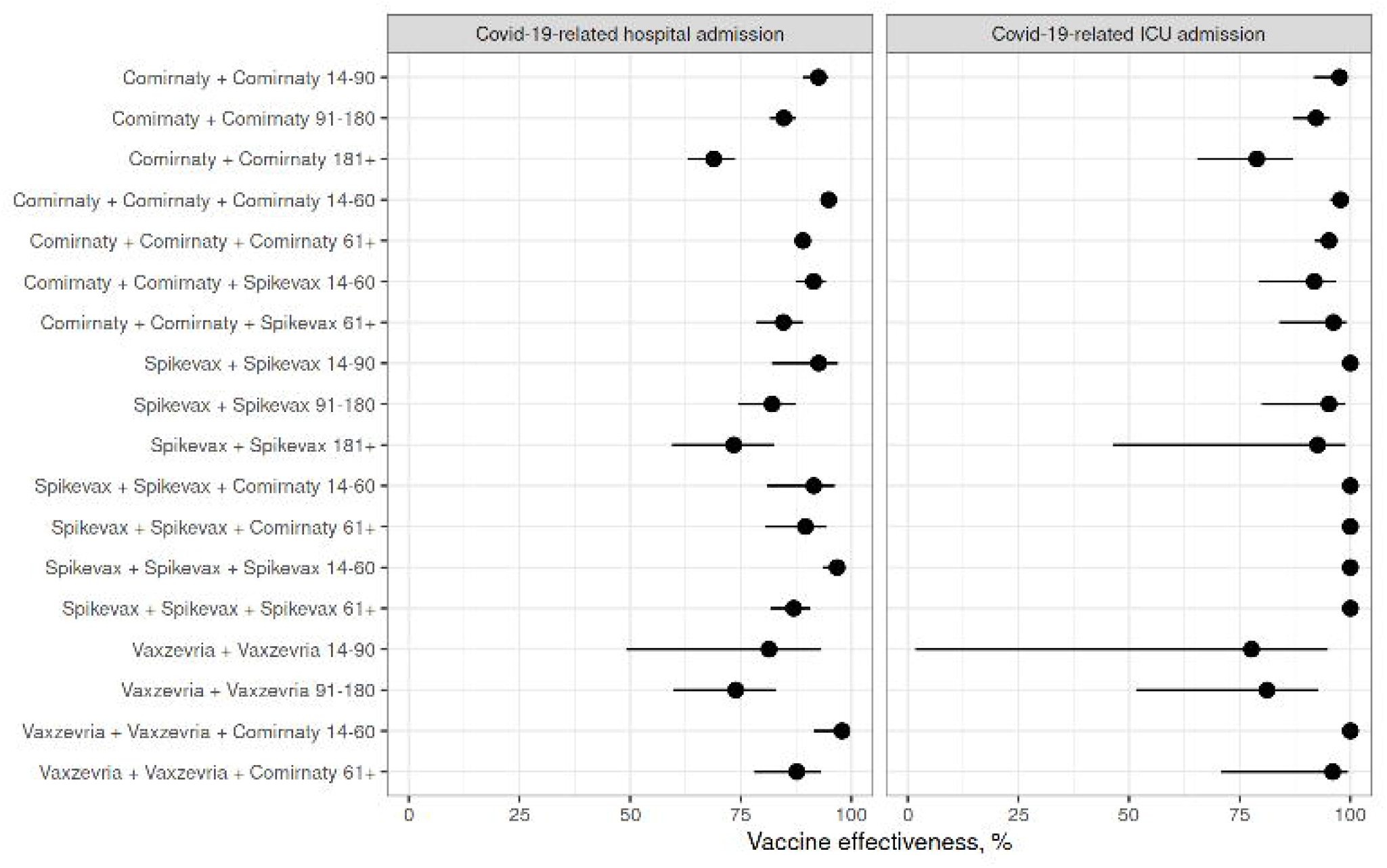
Vaccine effectiveness against primary outcomes by vaccine, dose and days since last vaccination. Footnote: Data points: point estimates; lines: 95% confidence interval estimates. All estimates are statistically significantly different from 0%.

Increasing age and comorbidities decreased the VE against hospitalization, yet the observed trend of waning remained unchanged (Supplementary Tables 10-11). In the 14–90 and 91–180 days after the second dose, the effectiveness of Comirnaty was 95% and 87% in 70-79-year-olds and 87% and 81% in 80-89-year-olds. In the 91–180 days after the second dose, the presence of either moderately or highly predisposing comorbidities reduced the VE from 92% to 83% or 82%. In the 14–60 days after the third dose of Comirnaty, the presence of comorbidities lowered the VE from 98% to 95% or 93%.

The effectiveness of Comirnaty was nearly constant over calendar time (Figure 2, Supplementary Figure 4). In the fourth quarter of 2021 (Q4), the VE against hospitalization was 90% (95% CI 78%–96%) in the first 14–90 days since the second vaccination and 96% (95% CI 93%–97%) in the first 14–60 days since the third vaccination (Figure 2, Supplementary Table 12). The corresponding estimates for the first quarter of 2022 (Q1), which was dominated by Omicron, were 91% (95% CI 83%–95%) and 94% (95% CI 92%–95%), respectively (Figure 2, Supplementary Table 13). However, while in Q4 there was almost no difference in point estimates in the 14–90 and 91–180 days after the second dose, in Q1 VE seemed to drop to 75% (95% CI 62%–83%) in the 91-180 days after the second dose (Figure 2). In both quarters, the median times since the second dose among those vaccinated 14–90 days ago (Q4: 63 days; Q1: 59 days) and 91–180 days ago (Q4: 146 days; Q1: 130 days) were similar.

**Figure 2.**
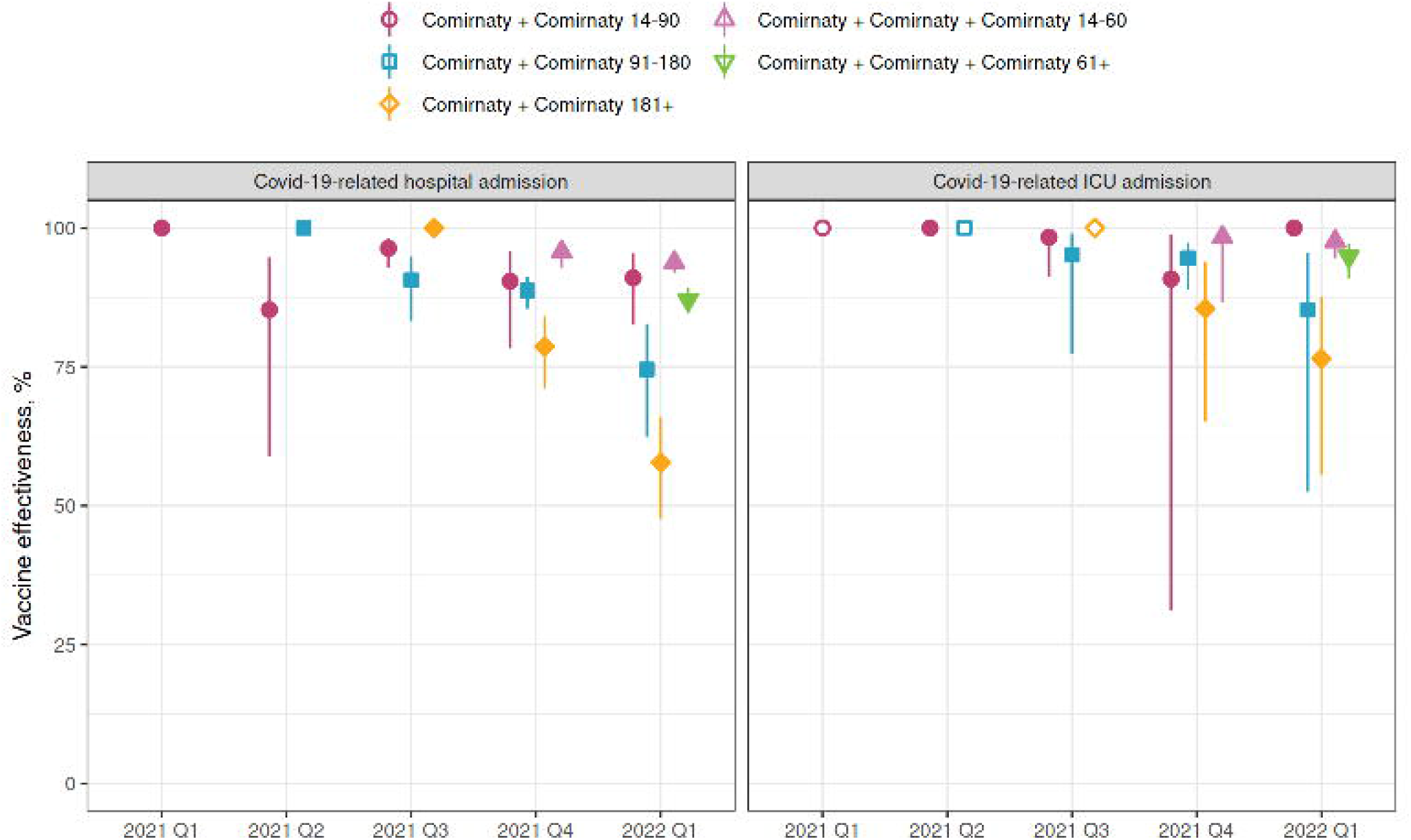
Comirnaty vaccine effectiveness against primary outcomes by quarter, dose and days since last vaccination. Footnote: Data points: point estimates; lines: 95% confidence interval estimates. A filled symbol indicates that the estimate is statistically significantly different from 0%, while an empty symbol indicates that the estimate is not statistically significantly different from 0%.

The control outcome, i.e., injuries, occurred 36 747 times in the study cohort. Without differentiating between vaccines, number of doses and time since vaccination, we estimated the hazard ratio at 0.96 (95% CI 0.93–1.00) and, thus, detected no statistically significant difference between the vaccinated and the unvaccinated. However, the hazard of injury in those vaccinated with Spikevax was higher than in the unvaccinated (Supplementary Table 14). The cumulative hazards in vaccinated and unvaccinated individuals were proportional over time (Supplementary Figure 5).

## Discussion

In this study of a nationwide elderly cohort, the Covid-19 vaccines used in Finland were highly effective against severe Covid-19. After the second and third dose, the VE against Covid-19-related hospitalization and ICU admission was over 90% for both Comirnaty and Spikevax. However, already during the first 6 months after the second dose, we observed signs of waning VE. Although the confidence intervals overlapped, VE against hospitalization appeared to decrease faster than VE against ICU admission. A third dose restored the level of protection to over 90%. Interestingly, the VE was nearly constant during the study period despite the sporadic emergence of new variants. However, the gradual drop in point estimates of VE against hospitalization was more evident in the first quarter of 2022, which might be an indicator of intensified waning after the second dose due to Omicron.

We found no meaningful difference in the effectiveness of the two mRNA vaccines. The effectiveness of Vaxzevria was, however, slightly lower, although it was still better than the average protection observed among the elderly after seasonal influenza vaccination (27,28). Nevertheless, individuals who were first vaccinated with two doses of Vaxzevria and later boosted with mRNA vaccine were as well protected as those who received three doses of mRNA vaccine.

The present study is concordant with other studies that were conducted prior to the emergence of Omicron and found limited waning of VE against Covid-19-related hospitalization during the first six months after the second dose in the elderly (4,7,13,14,29). In England, the VE against hospitalization decreased from initially excellent 98% to 91% within 5 months from the second vaccination with Comirnaty (4). As in the present study, Vaxzevria did not offer as high protection as Comirnaty and the VE was generally lower among individuals with chronic illnesses (4). In line with our findings, VE has also been reported to be reduced in the elderly aged 80 years and over compared to the younger elderly (14). Due to enhanced immunosenescence and illness- or treatment-induced immunosuppression, the protection offered by Covid-19 vaccines may, thus, be weaker among the very fragile elderly, such as residents of long-term facilities.

Recent studies of adult populations have estimated the VE against severe Covid-19 caused by Omicron at 50–90% after the second dose (16,21,24,25,30) and at approximately 90% after the third dose (16,21,25,30). Surprisingly, our results from the Omicron-dominated first quarter of 2022 match the upper limits of these estimates, although our study was restricted to elderly adults, in which VE is expected to be lower than that in younger adults (4,13,31). High VE against severe Covid-19 caused by Omicron may partially explain the relatively low hospital burden during the Omicron wave in many European countries despite skyrocketing infection rates (32). It is difficult to foresee how long the high level of protection will last. However, this study and another analysis (30) showed that the VE against severe Covid-19 remained at approximately 90% for at least 2–3 months after the third vaccination. The decision-making regarding the recommendation of a fourth dose for risk groups, such as the elderly, needs further evidence on the duration of natural and vaccine-induced immunity as well as VE against severe Covid-19 in various epidemiological settings.

Finland is one of the few countries that used an extended dosing interval of 12 weeks between the first and second dose due to shortage of Covid-19 vaccines in the early stage of the vaccination campaign (33). This interval is considered more immunogenic and might enhance VE against severe disease compared to the standard dosing interval of 3–4 weeks (7,34,35). Therefore, our results of higher VE might be explained by the longer dosing interval.

The present study has three major strengths. First, the nationwide cohort was highly representative as it covered essentially the whole elderly population in Finland. Second, the Finnish setting minimizes the risk of detection bias. Hospital care in Finland is equally accessible to all permanent residents due to the national health insurance resulting in low or no costs. Hospitalized patients with Covid-19 symptoms have been tested at low threshold irrespective of their vaccination status and the testing capacities in hospitals were at no point exhausted. Therefore, we assume that practically all elderly Covid-19 cases with severe symptoms requiring hospital care were identified as part of routine clinical practice and were thus eligible for inclusion in this study. Furthermore, we were able to exclude cases with concomitant asymptomatic or mild SARS-CoV-2 infection primarily hospitalized for other reasons than Covid-19. Third, we performed a negative control outcome analysis, which demonstrated that residual confounding after covariate adjustment would be negligible. Only those vaccinated with Spikevax had a statistically significantly higher risk of injuries than the unvaccinated indicating the presence of residual confounding due to differential behavior or frailty.

Although the national registers are known to be comprehensive and have been well maintained before and during the Covid-19 pandemic, the accuracy of the data has not been validated and information bias cannot be ruled out. Moreover, we assumed that the study population was randomly mixing so that vaccinated and unvaccinated subjects were equally exposed to the virus. Unfortunately, we have no data to support this assumption and differential vaccine uptake in particularly sheltered subpopulations such as residents of long-term care facilities may have thus led to bias. Although our findings may also apply to other populations of similar age, they do not necessarily apply to other outcomes. VE against mild disease or asymptomatic infection is likely lower than VE against severe disease. Another limitation is that the data do not allow estimation of variant-specific VE. As a surrogate we stratified the analysis by calendar time and used sequencing data to show which variant dominated each quarter.

## Conclusions

Since their introduction at the end of 2020, Covid-19 vaccines have been highly effective in preventing severe outcomes, such as Covid-19-related hospitalization and ICU admission, in the elderly, who carry the heaviest disease burden in a population. In our study, we observed signs of waning VE during the first six months after completion of a two-dose series, but a third dose restored the high level of protection for at least 2–3 months. Our analysis of data from the first quarter of 2022 suggests that the third dose still confers high protection against severe Covid-19 even after emergence of Omicron.

## Supporting information

Ethical concern

## Data Availability

By Finnish law, the authors are not permitted to share individual-level register data. The computing code is available upon request.

## Abbreviations

VE: Vaccine effectiveness
ICU: Intensive care unit
CI: Confidence interval
Q4: Fourth calendar quarter
Q1: First calendar quarter

## Declarations

### Ethics approval and consent to participate

By Finnish law, the Finnish Institute for Health and Welfare (THL) is the national expert institution to carry out surveillance on the impact of vaccinations in Finland (Communicable Diseases Act, https://www.finlex.fi/en/laki/kaannokset/2016/en20161227.pdf). Neither specific ethical approval of this study nor informed consent from the participants was needed.

### Consent for publication

Not applicable.

### Authors’ contributions

UB, EP, TL, TK, HN and AAP participated in the conceptualization of the study. UB conducted the statistical analysis and EP reviewed the literature. UB and EP drafted the manuscript. UB, EP, TL, TK, HN and AAP gave comments and revised the manuscript.

## Acknowledgments

The authors thank Heini Salo and Toni Lehtonen for the register-based identification of individuals with medical conditions predisposing to severe Covid-19 as well as Dorothée Obach, Eveline Otte im Kampe and Kari Auranen for their valuable comments during the conceptualization of the study (DO, EOiK) and manuscript revision (KA). Additional thanks go to all the colleagues at the Finnish Institute for Health and Welfare (THL) who curate the register data.

