## Supplement material for "High vaccine effectiveness against severe Covid-19 in the elderly in Finland before and after the emergence of Omicron"

### Supplementary Data

**Supplementary Table 1: Highly predisposing comorbidities or medical therapies** Definition of comorbidities and medical therapies that were considered highly predisposing to severe Covid-19.

| Classification system | Codes |
|---|---|
| **Actively treated cancer** | |
| ICD10 | C00, C01, C02, C03, C04, C05, C06, C07, C08, C09, C10, C11, C12, C13, C14, C15, C16, C17, C18, C19, C20, C21, C22, C23, C24, C25, C26, C27, C28, C29, C30, C31, C32, C33, C34, C35, C36, C37, C38, C39, C40, C41, C42, C43, C45, C46, C47, C48, C49, C50, C51, C52, C53, C54, C55, C56, C57, C58, C59, C60, C61, C62, C63, C64, C65, C66, C67, C68, C69, C70, C71, C72, C73, C74, C75, C76, C77, C78, C79, C80, C81, C82, C83, C84, C85, C86, C87, C88, C89, C90, C91, C92, C93, C94, C95, C96, C97, D051, D39 |
| **Down syndrome** | |
| ICD10 | Q90 |
| **Organ or stem cell transplantation** | |
| ICD10 | T86, Z94 |
| **Severe chronic respiratory disease** | |
| ICD10 | E84, J41, J42, J43, J44, J45, J46, J47, Z902 |
| ICPC2 | R96 |
| **Severe disorder of the immune system** | |
| ICD10 | D7081, D7089, D80, D81, D82, D83, D84, E3100, E723, E76, E791, E792, E798, E799, Q7782, Q7800, Q7808, Q809, Q8900, Q8933, Q8934, Q938, Z908 |
| NCSP | JMA |
| **Severe kidney disease** | |
| ICD10 | E102, E112, E142, I12, I13, N00, N01, N02, N03, N04, N05, N07, N08, N11, N14, N18, N19 |
| **Type 2 diabetes mellitus** | |
| ATC | A10B |
| ICD10 | E11, E13, E14 |
| ICPC2 | T90 |

ATC, Anatomical Therapeutic Chemical Classification System;

ICD10, International Statistical Classification of Diseases, tenth revision;

ICPC2, International Classification of Primary Care, second edition;

NCSP, Nordic Nomesco Classification of Surgical Procedures

**Supplementary Table 2: Moderately predisposing comorbidities or medical therapies** Definition of comorbidities and medical therapies that were considered moderately predisposing to severe Covid-19.

| Classification system | Codes |
|---|---|
| **Autoimmune disease** | |
| ICD10 | D86, K50, K51, L40, M02, M05, M06, M07, M139, M45, M460, M461, M469, M941 |
| **Immunosuppressive medication** | |
| ATC | H02AB02, H02AB04, H02AB06, H02AB07, L01BA01, L01XC02, L04AA06, L04AA10, L04AA13, L04AA18, L04AA24, L04AA26, L04AA29, L04AA33, L04AA37, L04AB, L04AC, L04AD01, L04AD02, L04AX01, L04AX03 |
| **Neurological condition affecting breathing** | |
| ICD10 | G20, G21, G22, G23, G24, G25, G26, G70, G71, G72, G73, G80, G81, G82, G83, I60, I61, I62, I63, I64, I65, I66, I67, I68, I69 |
| **Psychotic disease** | |
| ATC | N05AH02 |
| ICD10 | F20, F21, F22, F23, F24, F25, F26, F27, F28, F29 |
| ICPC2 | P72 |
| **Severe chronic liver disease** | |
| ICD10 | K702, K703, K704, K71, K72, K73, K74 |
| **Severe heart disease** | |
| ICD10 | I110, I119, I12, I130, I131, I132, I139, I15, I20, I21, I22, I23, I24, I25, I26, I27, I28, I418, I42, I43, I50 |
| **Sleep apnea** | |
| ICD10 | G473 |
| NCSP | WX723, WX780 |
| **Type 1 diabetes mellitus or adrenal insufficiency** | |
| ATC | A10A |
| ICD10 | E10, E250, E271, E272, E274, E3100, E3101, E3108, E896 |
| ICPC2 | T89 |

ATC, Anatomical Therapeutic Chemical Classification System;

ICD10, International Statistical Classification of Diseases, tenth revision;

ICPC2, International Classification of Primary Care, second edition;

NCSP, Nordic Nomesco Classification of Surgical Procedures

**Supplementary Table 3: Enrollment of the study cohort** Individuals were excluded if they had experienced any of the listed events prior to the start of the study.

| | Number of individuals |
|---|---|
| Target population | 898079 |
| Excluded due to prior laboratory-confirmed SARS-CoV-2 infection | 1856 |
| Excluded due to prior Covid-19-related hospital admission | <5 |
| Excluded due to prior Covid-19-related ICU admission | <5 |
| Excluded due to prior Covid-19 vaccination | <5 |
| Study cohort | 896220 |

**Supplementary Table 4: Distribution of baseline characteristics**

| | n | % |
|---|---|---|
| **Age in years** | | |
| 70-79 | 582456 | 65 |
| 80-89 | 257232 | 29 |
| 90-115 | 56532 | 6 |
| **Sex** | | |
| Male | 381794 | 43 |
| Female | 514426 | 57 |
| **Region of residence** | | |
| Helsinki-Uusimaa | 215597 | 24 |
| Åland | 4884 | 1 |
| Northern and Eastern Finland | 222845 | 25 |
| Southern Finland | 214011 | 24 |
| Western Finland | 238883 | 27 |
| **In long-term care** | | |
| No | 836441 | 93 |
| Yes | 59779 | 7 |
| **Influenza vaccination in 2019–2020** | | |
| No | 394840 | 44 |
| Yes | 501380 | 56 |
| **Nights hospitalized between 2015 and 2019** | | |
| 0 | 460923 | 51 |
| 1-5 | 191252 | 21 |
| 6-20 | 137503 | 15 |
| 21+ | 106542 | 12 |
| **Presence of comorbidities or medical therapies** | | |
| No predisposing comorbidities or medical therapies | 283025 | 32 |
| Only moderately predisposing comorbidities or medical therapies | 249467 | 28 |
| At least one highly predisposing comorbidity or medical therapy | 363728 | 41 |
| **Presence of particular comorbidities or medical therapies** | | |
| Severe heart disease | 366099 | 41 |
| Type 2 diabetes mellitus | 197819 | 22 |
| Immunosuppressive medication | 138701 | 15 |
| Actively treated cancer | 124874 | 14 |
| Severe chronic respiratory disease | 101222 | 11 |
| Neurological condition affecting breathing | 89043 | 10 |
| Autoimmune disease | 64897 | 7 |
| Type 1 diabetes mellitus or adrenal insufficiency | 52394 | 6 |
| Sleep apnea | 46101 | 5 |
| Severe kidney disease | 22440 | 3 |
| Psychotic disease | 11357 | 1 |
| Severe chronic liver disease | 4229 | 0 |
| Organ or stem cell transplantation | 2187 | 0 |
| Severe disorder of the immune system | 1555 | 0 |
| Down syndrome | 15 | 0 |

**Supplementary Table 5: Number of days until first dose** Distribution of the number of days until the receipt of the first dose in the study of vaccine effectiveness against Covid-19-related hospital admission.

| | Percentile | | |
|---|---|---|---|
| | 25th | 50th | 75th |
| **Age in years** | | | |
| 70-79 | 80 | 92 | 102 |
| 80-89 | 50 | 60 | 73 |
| 90-115 | 32 | 52 | 66 |
| **Sex** | | | |
| Male | 66 | 86 | 100 |
| Female | 60 | 80 | 94 |
| **Region of residence** | | | |
| Helsinki-Uusimaa | 59 | 80 | 93 |
| Åland | 48 | 69 | 84 |
| Northern and Eastern Finland | 62 | 82 | 101 |
| Southern Finland | 65 | 85 | 100 |
| Western Finland | 64 | 81 | 95 |
| **In long-term care** | | | |
| No | 66 | 84 | 97 |
| Yes | 23 | 30 | 47 |
| **Influenza vaccination in 2019–2020** | | | |
| No | 64 | 86 | 102 |
| Yes | 61 | 80 | 94 |
| **Nights hospitalized between 2015 and 2019** | | | |
| 0 | 68 | 87 | 101 |
| 1-5 | 65 | 81 | 95 |
| 6-20 | 57 | 74 | 93 |
| 21+ | 38 | 61 | 87 |
| **Presence of comorbidities or medical therapies** | | | |
| No predisposing comorbidities or medical therapies | 67 | 87 | 101 |
| Only moderately predisposing comorbidities or medical therapies | 59 | 80 | 94 |
| At least one highly predisposing comorbidity or medical therapy | 60 | 80 | 94 |

**Supplementary Table 6: Number of Covid-19-related hospital admissions** by vaccine, dose and days since last vaccination. The table shows the number of cases and person-years, the cumulative risk (per 100000) at the end of the study estimated based on the Kaplan-Meier estimator, the number of study subjects who contributed person-time, the number of study subjects who were censored before the end of the study and the number of study subjects who were still considered at risk at the end of the study. Study subjects may have passed through multiple exposure states before becoming cases or being censored.

| | Cases | P-years | Risk | Subjects | Censored | At risk |
|---|---|---|---|---|---|---|
| Not vaccinated | 793 | 244838 | 1313 | 896219 | 19439 | 34185 |
| Comirnaty 0-20 | 50 | 40761 | 951 | 709617 | 1274 | 144 |
| Comirnaty 21-83 | 84 | 114708 | 952 | 694046 | 6481 | 729 |
| Comirnaty 84+ | 34 | 10142 | 535 | 82994 | 5174 | 4257 |
| Comirnaty + Comirnaty 0-13 | 9 | 26489 | 442 | 691558 | 636 | 408 |
| Comirnaty + Comirnaty 14-90 | 33 | 144412 | 108 | 690508 | 8215 | 4065 |
| Comirnaty + Comirnaty 91-180 | 210 | 158116 | 277 | 677291 | 9830 | 14194 |
| Comirnaty + Comirnaty 181+ | 200 | 31354 | 457 | 503537 | 7918 | 24383 |
| Comirnaty + Comirnaty + Comirnaty 0-13 | 40 | 20968 | 186 | 548987 | 663 | 2492 |
| Comirnaty + Comirnaty + Comirnaty 14-60 | 107 | 69468 | 84 | 545832 | 7024 | 8223 |
| Comirnaty + Comirnaty + Comirnaty 61+ | 359 | 74262 | 125 | 530585 | 48349 | 482236 |
| Comirnaty + Comirnaty + Spikevax 0-13 | 10 | 2787 | 69 | 72922 | 73 | 279 |
| Comirnaty + Comirnaty + Spikevax 14-60 | 28 | 9222 | 118 | 72570 | 723 | 2907 |
| Comirnaty + Comirnaty + Spikevax 61+ | 39 | 5601 | 325 | 68940 | 1674 | 67266 |
| Spikevax 0-20 | <5 | 4901 | 386 | 85310 | 171 | 13 |
| Spikevax 21-83 | 10 | 13902 | 466 | 85094 | 2470 | 187 |
| Spikevax 84+ | 8 | 1662 | 825 | 13902 | 1917 | 851 |
| Spikevax + Spikevax 0-13 | <5 | 3053 | 9 | 79701 | 77 | 62 |
| Spikevax + Spikevax 14-90 | 5 | 16589 | 201 | 79561 | 1126 | 812 |
| Spikevax + Spikevax 91-180 | 34 | 17796 | 277 | 77410 | 1319 | 2308 |
| Spikevax + Spikevax 181+ | 23 | 3315 | 433 | 53206 | 784 | 3232 |
| Spikevax + Spikevax + Comirnaty 0-13 | <5 | 666 | 175 | 17504 | 38 | 164 |
| Spikevax + Spikevax + Comirnaty 14-60 | 6 | 2167 | 137 | 17302 | 374 | 666 |
| Spikevax + Spikevax + Comirnaty 61+ | 10 | 2092 | 70 | 16262 | 1314 | 14948 |
| Spikevax + Spikevax + Spikevax 0-13 | 5 | 2006 | 70 | 52477 | 75 | 169 |
| Spikevax + Spikevax + Spikevax 14-60 | 8 | 6630 | 27 | 52233 | 746 | 1113 |
| Spikevax + Spikevax + Spikevax 61+ | 41 | 5948 | 109 | 50374 | 2585 | 47789 |
| Vaxzevria 0-20 | <5 | 2740 | 14 | 47669 | 45 | 0 |
| Vaxzevria 21-83 | <5 | 7965 | 7 | 47612 | 5404 | 0 |
| Vaxzevria 84+ | <5 | 474 | 342 | 5100 | 2812 | 172 |
| Vaxzevria + Vaxzevria 0-13 | 0 | 1504 | 0 | 39236 | 17 | 0 |
| Vaxzevria + Vaxzevria 14-90 | <5 | 8248 | 16 | 39219 | 208 | 0 |
| Vaxzevria + Vaxzevria 91-180 | 23 | 9067 | 205 | 38962 | 400 | 6 |
| Vaxzevria + Vaxzevria 181+ | 18 | 1283 | 379 | 24663 | 319 | 1896 |
| Vaxzevria + Vaxzevria + Comirnaty 0-13 | <5 | 919 | 49 | 24014 | 19 | 74 |
| Vaxzevria + Vaxzevria + Comirnaty 14-60 | <5 | 3044 | 24 | 23921 | 224 | 669 |
| Vaxzevria + Vaxzevria + Comirnaty 61+ | 12 | 2434 | 363 | 23028 | 468 | 22560 |
| Vaxzevria + Vaxzevria + Spikevax 0-13 | <5 | 474 | 31 | 12376 | 7 | 12 |
| Vaxzevria + Vaxzevria + Spikevax 14-60 | <5 | 1573 | 95 | 12357 | 79 | 536 |
| Vaxzevria + Vaxzevria + Spikevax 61+ | 5 | 939 | 102 | 11742 | 111 | 11631 |

**Supplementary Table 7: Number of Covid-19-related ICU admissions** by vaccine, dose and days since last vaccination. The table shows the number of cases and person-years, the cumulative risk (per 100000) at the end of the study estimated based on the Kaplan-Meier estimator, the number of study subjects who contributed person-time, the number of study subjects who were censored before the end of the study and the number of study subjects who were still considered at risk at the end of the study. Study subjects may have passed through multiple exposure states before becoming cases or being censored.

| | Cases | P-years | Risk | Subjects | Censored | At risk |
|---:|---:|---:|---:|---:|---:|---:|
| Not vaccinated | 159 | 244856 | 238 | 896219 | 19408 | 34215 |
| Comirnaty 0-20 | 10 | 40762 | 75 | 709618 | 1246 | 144 |
| Comirnaty 21-83 | 8 | 114711 | 14 | 694075 | 6496 | 729 |
| Comirnaty 84+ | <5 | 10143 | 68 | 83006 | 5184 | 4259 |
| Comirnaty + Comirnaty 0-13 | <5 | 26489 | 2 | 691560 | 630 | 408 |
| Comirnaty + Comirnaty 14-90 | <5 | 144413 | 8 | 690516 | 8212 | 4065 |
| Comirnaty + Comirnaty 91-180 | 26 | 158121 | 29 | 677301 | 9788 | 14199 |
| Comirnaty + Comirnaty 181+ | 22 | 31360 | 55 | 503584 | 7954 | 24393 |
| Comirnaty + Comirnaty + Comirnaty 0-13 | <5 | 20969 | 2 | 548989 | 626 | 2492 |
| Comirnaty + Comirnaty + Comirnaty 14-60 | 9 | 69472 | 3 | 545871 | 7021 | 8227 |
| Comirnaty + Comirnaty + Comirnaty 61+ | 26 | 74271 | 55 | 530623 | 48271 | 482352 |
| Comirnaty + Comirnaty + Spikevax 0-13 | <5 | 2787 | 15 | 72922 | 65 | 279 |
| Comirnaty + Comirnaty + Spikevax 14-60 | 5 | 9222 | 13 | 72578 | 724 | 2907 |
| Comirnaty + Comirnaty + Spikevax 61+ | <5 | 5602 | 4 | 68947 | 1671 | 67276 |
| Spikevax 0-20 | 0 | 4901 | 0 | 85310 | 168 | 13 |
| Spikevax 21-83 | <5 | 13902 | 4 | 85097 | 2473 | 187 |
| Spikevax 84+ | 0 | 1662 | 0 | 13902 | 1915 | 853 |
| Spikevax + Spikevax 0-13 | 0 | 3053 | 0 | 79701 | 76 | 62 |
| Spikevax + Spikevax 14-90 | 0 | 16589 | 0 | 79562 | 1125 | 812 |
| Spikevax + Spikevax 91-180 | <5 | 17797 | 45 | 77412 | 1317 | 2308 |
| Spikevax + Spikevax 181+ | <5 | 3316 | 30 | 53210 | 784 | 3235 |
| Spikevax + Spikevax + Comirnaty 0-13 | 0 | 666 | 0 | 17504 | 35 | 164 |
| Spikevax + Spikevax + Comirnaty 14-60 | 0 | 2167 | 0 | 17305 | 376 | 666 |
| Spikevax + Spikevax + Comirnaty 61+ | 0 | 2092 | 0 | 16263 | 1309 | 14954 |
| Spikevax + Spikevax + Spikevax 0-13 | 0 | 2007 | 0 | 52478 | 70 | 170 |
| Spikevax + Spikevax + Spikevax 14-60 | 0 | 6630 | 0 | 52238 | 749 | 1113 |
| Spikevax + Spikevax + Spikevax 61+ | 0 | 5949 | 0 | 50376 | 2573 | 47803 |
| Vaxzevria 0-20 | 0 | 2740 | 0 | 47669 | 42 | 0 |
| Vaxzevria 21-83 | <5 | 7965 | 4167 | 47615 | 5408 | 0 |
| Vaxzevria 84+ | 0 | 474 | 0 | 5099 | 2811 | 172 |
| Vaxzevria + Vaxzevria 0-13 | 0 | 1504 | 0 | 39236 | 17 | 0 |
| Vaxzevria + Vaxzevria 14-90 | <5 | 8248 | 10 | 39219 | 208 | 0 |
| Vaxzevria + Vaxzevria 91-180 | 5 | 9067 | 13 | 38962 | 397 | 6 |
| Vaxzevria + Vaxzevria 181+ | <5 | 1283 | 71 | 24666 | 322 | 1896 |
| Vaxzevria + Vaxzevria + Comirnaty 0-13 | 0 | 919 | 0 | 24014 | 17 | 74 |
| Vaxzevria + Vaxzevria + Comirnaty 14-60 | 0 | 3044 | 0 | 23923 | 226 | 669 |
| Vaxzevria + Vaxzevria + Comirnaty 61+ | <5 | 2434 | 307 | 23028 | 462 | 22566 |
| Vaxzevria + Vaxzevria + Spikevax 0-13 | 0 | 474 | 0 | 12376 | 6 | 12 |
| Vaxzevria + Vaxzevria + Spikevax 14-60 | 0 | 1573 | 0 | 12358 | 77 | 536 |
| Vaxzevria + Vaxzevria + Spikevax 61+ | <5 | 939 | 11 | 11745 | 111 | 11634 |

**Supplementary Table 8: VE against Covid-19-related hospital admission** Vaccine effectiveness (in %) quantified as one minus the hazard ratio adjusted for age, sex, region of residence, residence in a long-term care facility, influenza vaccination in 2019–2020, number of nights hospitalized between 2015 and 2019 and presence of predisposing comorbidities or medical therapies.

| | MLE | LCI | UCI | p-value[1] |
|---:|---:|---:|---:|---:|
| Comirnaty 0-20 | 45 | 26 | 60 | . |
| Comirnaty 21-83 | 61 | 49 | 71 | . |
| Comirnaty 84+ | 64 | 49 | 75 | . |
| Comirnaty + Comirnaty 0-13 | 80 | 61 | 90 | . |
| Comirnaty + Comirnaty 14-90 | 93 | 89 | 95 | . |
| Comirnaty + Comirnaty 91-180 | 85 | 82 | 87 | . |
| Comirnaty + Comirnaty 181+ | 69 | 63 | 74 | . |
| Comirnaty + Comirnaty + Comirnaty 0-13 | 89 | 85 | 92 | . |
| Comirnaty + Comirnaty + Comirnaty 14-60 | 95 | 94 | 96 | . |
| Comirnaty + Comirnaty + Comirnaty 61+ | 89 | 87 | 91 | . |
| Comirnaty + Comirnaty + Spikevax 0-13 | 87 | 75 | 93 | . |
| Comirnaty + Comirnaty + Spikevax 14-60 | 91 | 87 | 94 | . |
| Comirnaty + Comirnaty + Spikevax 61+ | 85 | 78 | 89 | . |
| Spikevax 0-20 | 72 | 24 | 90 | . |
| Spikevax 21-83 | 70 | 43 | 84 | . |
| Spikevax 84+ | 63 | 26 | 82 | . |
| Spikevax + Spikevax 0-13 | 86 | 0 | 98 | . |
| Spikevax + Spikevax 14-90 | 93 | 82 | 97 | . |
| Spikevax + Spikevax 91-180 | 82 | 74 | 87 | . |
| Spikevax + Spikevax 181+ | 73 | 59 | 83 | . |
| Spikevax + Spikevax + Comirnaty 0-13 | 78 | 32 | 93 | . |
| Spikevax + Spikevax + Comirnaty 14-60 | 91 | 81 | 96 | . |
| Spikevax + Spikevax + Comirnaty 61+ | 90 | 80 | 94 | . |
| Spikevax + Spikevax + Spikevax 0-13 | 90 | 75 | 96 | . |
| Spikevax + Spikevax + Spikevax 14-60 | 97 | 93 | 98 | . |
| Spikevax + Spikevax + Spikevax 61+ | 87 | 82 | 91 | . |
| Vaxzevria 0-20 | 32 | -116 | 78 | . |
| Vaxzevria 21-83 | 68 | -3 | 90 | . |
| Vaxzevria 84+ | 37 | -95 | 80 | . |
| Vaxzevria + Vaxzevria 0-13 | 100 | . | . | 0.132 |
| Vaxzevria + Vaxzevria 14-90 | 81 | 49 | 93 | . |
| Vaxzevria + Vaxzevria 91-180 | 74 | 60 | 83 | . |
| Vaxzevria + Vaxzevria 181+ | 48 | 17 | 68 | . |
| Vaxzevria + Vaxzevria + Comirnaty 0-13 | 84 | 51 | 95 | . |
| Vaxzevria + Vaxzevria + Comirnaty 14-60 | 98 | 91 | 99 | . |
| Vaxzevria + Vaxzevria + Comirnaty 61+ | 88 | 78 | 93 | . |
| Vaxzevria + Vaxzevria + Spikevax 0-13 | 92 | 41 | 99 | . |
| Vaxzevria + Vaxzevria + Spikevax 14-60 | 94 | 82 | 98 | . |
| Vaxzevria + Vaxzevria + Spikevax 61+ | 87 | 70 | 95 | . |

MLE, maximum likelihood estimate;

LCI/UCI, lower/upper limit of the 95% Wald confidence interval

[1] Likelihood-ratio test

**Supplementary Table 9: VE against Covid-19-related ICU admission** Vaccine effectiveness (in %) quantified as one minus the hazard ratio adjusted for age, sex, region of residence, residence in a long-term care facility, influenza vaccination in 2019–2020, number of nights hospitalized between 2015 and 2019 and presence of predisposing comorbidities or medical therapies.

| | MLE | LCI | UCI | p-value[1] |
|---:|---:|---:|---:|---:|
| Comirnaty 0-20 | 57 | 13 | 79 | . |
| Comirnaty 21-83 | 84 | 63 | 93 | . |
| Comirnaty 84+ | 69 | 15 | 89 | . |
| Comirnaty + Comirnaty 0-13 | 75 | -10 | 94 | . |
| Comirnaty + Comirnaty 14-90 | 98 | 92 | 99 | . |
| Comirnaty + Comirnaty 91-180 | 92 | 87 | 95 | . |
| Comirnaty + Comirnaty 181+ | 79 | 65 | 87 | . |
| Comirnaty + Comirnaty + Comirnaty 0-13 | 99 | 91 | 100 | . |
| Comirnaty + Comirnaty + Comirnaty 14-60 | 98 | 95 | 99 | . |
| Comirnaty + Comirnaty + Comirnaty 61+ | 95 | 92 | 97 | . |
| Comirnaty + Comirnaty + Spikevax 0-13 | 94 | 54 | 99 | . |
| Comirnaty + Comirnaty + Spikevax 14-60 | 92 | 79 | 97 | . |
| Comirnaty + Comirnaty + Spikevax 61+ | 96 | 84 | 99 | . |
| Spikevax 0-20 | 100 | . | . | 0.016 |
| Spikevax 21-83 | 70 | -29 | 93 | . |
| Spikevax 84+ | 100 | . | . | 0.018 |
| Spikevax + Spikevax 0-13 | 100 | . | . | 0.122 |
| Spikevax + Spikevax 14-90 | 100 | . | . | <0.001 |
| Spikevax + Spikevax 91-180 | 95 | 80 | 99 | . |
| Spikevax + Spikevax 181+ | 93 | 46 | 99 | . |
| Spikevax + Spikevax + Comirnaty 0-13 | 100 | . | . | 0.022 |
| Spikevax + Spikevax + Comirnaty 14-60 | 100 | . | . | <0.001 |
| Spikevax + Spikevax + Comirnaty 61+ | 100 | . | . | <0.001 |
| Spikevax + Spikevax + Spikevax 0-13 | 100 | . | . | <0.001 |
| Spikevax + Spikevax + Spikevax 14-60 | 100 | . | . | <0.001 |
| Spikevax + Spikevax + Spikevax 61+ | 100 | . | . | <0.001 |
| Vaxzevria 0-20 | 100 | . | . | 0.074 |
| Vaxzevria 21-83 | 68 | -151 | 96 | . |
| Vaxzevria 84+ | 100 | . | . | 0.140 |
| Vaxzevria + Vaxzevria 0-13 | 100 | . | . | 0.509 |
| Vaxzevria + Vaxzevria 14-90 | 78 | 2 | 95 | . |
| Vaxzevria + Vaxzevria 91-180 | 81 | 52 | 93 | . |
| Vaxzevria + Vaxzevria 181+ | 64 | -14 | 89 | . |
| Vaxzevria + Vaxzevria + Comirnaty 0-13 | 100 | . | . | 0.002 |
| Vaxzevria + Vaxzevria + Comirnaty 14-60 | 100 | . | . | <0.001 |
| Vaxzevria + Vaxzevria + Comirnaty 61+ | 96 | 71 | 99 | . |
| Vaxzevria + Vaxzevria + Spikevax 0-13 | 100 | . | . | 0.021 |
| Vaxzevria + Vaxzevria + Spikevax 14-60 | 100 | . | . | <0.001 |
| Vaxzevria + Vaxzevria + Spikevax 61+ | 89 | 22 | 99 | . |

MLE, maximum likelihood estimate;

LCI/UCI, lower/upper limit of the 95% Wald confidence interval

[1] Likelihood-ratio test

**Supplementary Table 10: VE against Covid-19-related hospital admission by age group** Vaccine effectiveness (in %) quantified as one minus the hazard ratio stratified by age in years and adjusted for sex, region of residence, residence in a long-term care facility, influenza vaccination in 2019–2020, number of nights hospitalized between 2015 and 2019 and presence of predisposing comorbidities or medical therapies. Estimates marked by an asterisk are statistically significantly different from the corresponding estimates for 70–79-year-olds.

| | 70-79 | 80-89 | 90-115 |
|---:|---:|---:|---:|
| Comirnaty 0-20 | 41 | 44 | 46 |
| Comirnaty 21-83 | 67 | 47* | 62 |
| Comirnaty 84+ | 66 | 66 | 31 |
| Comirnaty + Comirnaty 0-13 | 87 | 66 | 100 |
| Comirnaty + Comirnaty 14-90 | 95 | 87* | 89 |
| Comirnaty + Comirnaty 91-180 | 87 | 81* | 88 |
| Comirnaty + Comirnaty 181+ | 73 | 65 | 48* |
| Comirnaty + Comirnaty + Comirnaty 0-13 | 91 | 90 | 67* |
| Comirnaty + Comirnaty + Comirnaty 14-60 | 96 | 94 | 91 |
| Comirnaty + Comirnaty + Comirnaty 61+ | 91 | 88 | 79* |
| Comirnaty + Comirnaty + Spikevax 0-13 | 92 | 69* | 100 |
| Comirnaty + Comirnaty + Spikevax 14-60 | 94 | 87 | 74 |
| Comirnaty + Comirnaty + Spikevax 61+ | 89 | 81 | 24* |
| Spikevax 0-20 | 58 | 100 | 2 |
| Spikevax 21-83 | 76 | 83 | -61* |
| Spikevax 84+ | 89 | 35 | 53 |
| Spikevax + Spikevax 0-13 | 100 | 61 | 100 |
| Spikevax + Spikevax 14-90 | 98 | 81 | 100 |
| Spikevax + Spikevax 91-180 | 87 | 74 | 70 |
| Spikevax + Spikevax 181+ | 79 | 68 | 57 |
| Spikevax + Spikevax + Comirnaty 0-13 | 100 | 78 | -187 |
| Spikevax + Spikevax + Comirnaty 14-60 | 93 | 88 | 100 |
| Spikevax + Spikevax + Comirnaty 61+ | 94 | 82 | 100 |
| Spikevax + Spikevax + Spikevax 0-13 | 91 | 93 | 62 |
| Spikevax + Spikevax + Spikevax 14-60 | 99 | 95 | 86* |
| Spikevax + Spikevax + Spikevax 61+ | 89 | 87 | 73 |
| Vaxzevria 0-20 | 28 | 100 | 100 |
| Vaxzevria 21-83 | 68 | 100 | 100 |
| Vaxzevria 84+ | 52 | -29 | 100 |
| Vaxzevria + Vaxzevria 0-13 | 100 | 100 | 100 |
| Vaxzevria + Vaxzevria 14-90 | 87 | 22 | 100 |
| Vaxzevria + Vaxzevria 91-180 | 75 | 100 | 100 |
| Vaxzevria + Vaxzevria 181+ | 50 | 68 | 100 |
| Vaxzevria + Vaxzevria + Comirnaty 0-13 | 85 | 100 | 100 |
| Vaxzevria + Vaxzevria + Comirnaty 14-60 | 98 | 100 | 100 |
| Vaxzevria + Vaxzevria + Comirnaty 61+ | 89 | 76 | 100 |
| Vaxzevria + Vaxzevria + Spikevax 0-13 | 92 | 100 | 100 |
| Vaxzevria + Vaxzevria + Spikevax 14-60 | 95 | 100 | 100 |
| Vaxzevria + Vaxzevria + Spikevax 61+ | 88 | 100 | 100 |

**Supplementary Table 11: VE against Covid-19-related hospital admission by presence of comorbidities** Vaccine effectiveness (in %) quantified as one minus the hazard ratio stratified by presence of predisposing comorbidities or medical therapies and adjusted for age, sex, region of residence, residence in a long-term care facility, influenza vaccination in 2019–2020 and number of nights hospitalized between 2015 and 2019. Estimates marked by an asterisk are statistically significantly different from the corresponding estimates for individuals without predisposing comorbidities or medical therapies.

| | None | Moderate | High |
|---|---|---|---|
| Comirnaty 0-20 | 56 | 43 | 41 |
| Comirnaty 21-83 | 54 | 82* | 53 |
| Comirnaty 84+ | 57 | 64 | 64 |
| Comirnaty + Comirnaty 0-13 | 88 | 100 | 67 |
| Comirnaty + Comirnaty 14-90 | 96 | 90 | 92 |
| Comirnaty + Comirnaty 91-180 | 92 | 83* | 82* |
| Comirnaty + Comirnaty 181+ | 80 | 68 | 63* |
| Comirnaty + Comirnaty + Comirnaty 0-13 | 96 | 89 | 86* |
| Comirnaty + Comirnaty + Comirnaty 14-60 | 98 | 95* | 93* |
| Comirnaty + Comirnaty + Comirnaty 61+ | 95 | 87* | 87* |
| Comirnaty + Comirnaty + Spikevax 0-13 | 94 | 94 | 80 |
| Comirnaty + Comirnaty + Spikevax 14-60 | 97 | 93 | 88* |
| Comirnaty + Comirnaty + Spikevax 61+ | 98 | 78* | 82* |
| Spikevax 0-20 | 54 | 100 | 64 |
| Spikevax 21-83 | 80 | 87 | 58 |
| Spikevax 84+ | 100 | 62 | 52 |
| Spikevax + Spikevax 0-13 | 100 | 100 | 75 |
| Spikevax + Spikevax 14-90 | 91 | 87 | 95 |
| Spikevax + Spikevax 91-180 | 97 | 77 | 78 |
| Spikevax + Spikevax 181+ | 85 | 66 | 71 |
| Spikevax + Spikevax + Comirnaty 0-13 | 100 | 68 | 74 |
| Spikevax + Spikevax + Comirnaty 14-60 | 92 | 88 | 92 |
| Spikevax + Spikevax + Comirnaty 61+ | 100 | 100 | 82 |
| Spikevax + Spikevax + Spikevax 0-13 | 100 | 91 | 85 |
| Spikevax + Spikevax + Spikevax 14-60 | 100 | 98 | 95 |
| Spikevax + Spikevax + Spikevax 61+ | 94 | 85 | 85 |
| Vaxzevria 0-20 | 1 | 100 | 17 |
| Vaxzevria 21-83 | 100 | -4 | 80 |
| Vaxzevria 84+ | -110 | -1 | 100 |
| Vaxzevria + Vaxzevria 0-13 | 100 | 100 | 100 |
| Vaxzevria + Vaxzevria 14-90 | 100 | 100 | 65 |
| Vaxzevria + Vaxzevria 91-180 | 81 | 95 | 61 |
| Vaxzevria + Vaxzevria 181+ | 88 | 73 | 19 |
| Vaxzevria + Vaxzevria + Comirnaty 0-13 | 100 | 100 | 70 |
| Vaxzevria + Vaxzevria + Comirnaty 14-60 | 100 | 90 | 100 |
| Vaxzevria + Vaxzevria + Comirnaty 61+ | 100 | 95 | 79 |
| Vaxzevria + Vaxzevria + Spikevax 0-13 | 67 | 100 | 100 |
| Vaxzevria + Vaxzevria + Spikevax 14-60 | 93 | 91 | 96 |
| Vaxzevria + Vaxzevria + Spikevax 61+ | 80 | 88 | 90 |

**Supplementary Table 12: VE against Covid-19-related hospital admission in 2021 Q4**, i.e., between October 1 and December 31. Vaccine effectiveness (in %) quantified as one minus the hazard ratio adjusted for age, sex, region of residence, residence in a long-term care facility, influenza vaccination in 2019–2020, number of nights hospitalized between 2015 and 2019 and presence of predisposing comorbidities or medical therapies.

| | Cases | P-years | MLE | LCI | UCI | p-value[1] |
|---|---|---|---|---|---|---|
| Not vaccinated | 150 | 11015 | . | . | . | . |
| Comirnaty 0-20 | <5 | 411 | 65 | -40 | 91 | . |
| Comirnaty 21-83 | 5 | 773 | 53 | -16 | 81 | . |
| Comirnaty 84+ | 15 | 3312 | 68 | 45 | 81 | . |
| Comirnaty + Comirnaty 0-13 | <5 | 754 | 83 | 31 | 96 | . |
| Comirnaty + Comirnaty 14-90 | 6 | 4694 | 90 | 78 | 96 | . |
| Comirnaty + Comirnaty 91-180 | 157 | 103650 | 89 | 85 | 91 | . |
| Comirnaty + Comirnaty 181+ | 76 | 19396 | 79 | 71 | 84 | . |
| Comirnaty + Comirnaty + Comirnaty 0-13 | 23 | 16526 | 93 | 88 | 95 | . |
| Comirnaty + Comirnaty + Comirnaty 14-60 | 18 | 18309 | 96 | 93 | 97 | . |
| Comirnaty + Comirnaty + Comirnaty 61+ | <5 | 1308 | 94 | 75 | 99 | . |
| Comirnaty + Comirnaty + Spikevax 0-13 | 0 | 808 | 100 | . | . | <0.001 |
| Comirnaty + Comirnaty + Spikevax 14-60 | <5 | 354 | 75 | -3 | 94 | . |
| Comirnaty + Comirnaty + Spikevax 61+ | 0 | 100 | 100 | . | . | 0.021 |
| Spikevax 0-20 | 0 | 73 | 100 | . | . | 0.141 |
| Spikevax 21-83 | <5 | 196 | 60 | -189 | 94 | . |
| Spikevax 84+ | <5 | 621 | 91 | 35 | 99 | . |
| Spikevax + Spikevax 0-13 | 0 | 129 | 100 | . | . | 0.038 |
| Spikevax + Spikevax 14-90 | <5 | 862 | 92 | 42 | 99 | . |
| Spikevax + Spikevax 91-180 | 22 | 12397 | 88 | 81 | 93 | . |
| Spikevax + Spikevax 181+ | 6 | 2085 | 87 | 70 | 94 | . |
| Spikevax + Spikevax + Comirnaty 0-13 | <5 | 481 | 79 | 13 | 95 | . |
| Spikevax + Spikevax + Comirnaty 14-60 | <5 | 458 | 82 | 28 | 96 | . |
| Spikevax + Spikevax + Comirnaty 61+ | 0 | 15 | 100 | . | . | 0.304 |
| Spikevax + Spikevax + Spikevax 0-13 | <5 | 1288 | 93 | 73 | 98 | . |
| Spikevax + Spikevax + Spikevax 14-60 | 0 | 1148 | 100 | . | . | <0.001 |
| Spikevax + Spikevax + Spikevax 61+ | 0 | 29 | 100 | . | . | 0.145 |
| Vaxzevria 0-20 | 0 | <5 | 100 | . | . | 0.953 |
| Vaxzevria 21-83 | 0 | <5 | 100 | . | . | 0.807 |
| Vaxzevria 84+ | <5 | 192 | 62 | -171 | 95 | . |
| Vaxzevria + Vaxzevria 0-13 | 0 | <5 | 100 | . | . | 0.894 |
| Vaxzevria + Vaxzevria 14-90 | 0 | 295 | 100 | . | . | 0.023 |
| Vaxzevria + Vaxzevria 91-180 | 21 | 8040 | 79 | 67 | 87 | . |
| Vaxzevria + Vaxzevria 181+ | 6 | 411 | 23 | -76 | 67 | . |
| Vaxzevria + Vaxzevria + Comirnaty 0-13 | <5 | 528 | 90 | 26 | 99 | . |
| Vaxzevria + Vaxzevria + Comirnaty 14-60 | <5 | 283 | 84 | -16 | 98 | . |
| Vaxzevria + Vaxzevria + Comirnaty 61+ | 0 | 14 | 100 | . | . | 0.365 |
| Vaxzevria + Vaxzevria + Spikevax 0-13 | 0 | 156 | 100 | . | . | 0.012 |
| Vaxzevria + Vaxzevria + Spikevax 14-60 | 0 | 42 | 100 | . | . | 0.171 |
| Vaxzevria + Vaxzevria + Spikevax 61+ | 0 | <5 | 100 | . | . | 0.695 |

MLE, maximum likelihood estimate;

LCI/UCI, lower/upper limit of the 95% Wald confidence interval

[1] Likelihood-ratio test

**Supplementary Table 13: VE against Covid-19-related hospital admission in 2022 Q1**, i.e., between January 1 and March 31. Vaccine effectiveness (in %) quantified as one minus the hazard ratio adjusted for age, sex, region of residence, residence in a long-term care facility, influenza vaccination in 2019–2020, number of nights hospitalized between 2015 and 2019 and presence of predisposing comorbidities or medical therapies.

| | Cases | P-years | MLE | LCI | UCI | p-value[1] |
|---|---|---|---|---|---|---|
| Not vaccinated | 292 | 8929 | . | . | . | . |
| Comirnaty 0-20 | <5 | 122 | 24 | -137 | 76 | . |
| Comirnaty 21-83 | 12 | 460 | 22 | -38 | 56 | . |
| Comirnaty 84+ | 15 | 1073 | 61 | 34 | 77 | . |
| Comirnaty + Comirnaty 0-13 | <5 | 200 | 67 | -31 | 92 | . |
| Comirnaty + Comirnaty 14-90 | 9 | 2993 | 91 | 83 | 95 | . |
| Comirnaty + Comirnaty 91-180 | 28 | 3323 | 75 | 62 | 83 | . |
| Comirnaty + Comirnaty 181+ | 124 | 9161 | 58 | 48 | 66 | . |
| Comirnaty + Comirnaty + Comirnaty 0-13 | 17 | 4384 | 84 | 73 | 90 | . |
| Comirnaty + Comirnaty + Comirnaty 14-60 | 89 | 51159 | 94 | 92 | 95 | . |
| Comirnaty + Comirnaty + Comirnaty 61+ | 357 | 72954 | 87 | 85 | 89 | . |
| Comirnaty + Comirnaty + Spikevax 0-13 | 10 | 1963 | 82 | 66 | 91 | . |
| Comirnaty + Comirnaty + Spikevax 14-60 | 26 | 8868 | 91 | 87 | 94 | . |
| Comirnaty + Comirnaty + Spikevax 61+ | 39 | 5501 | 83 | 76 | 88 | . |
| Spikevax 0-20 | <5 | 42 | 38 | -340 | 91 | . |
| Spikevax 21-83 | <5 | 122 | 81 | -33 | 97 | . |
| Spikevax 84+ | 7 | 204 | 23 | -63 | 64 | . |
| Spikevax + Spikevax 0-13 | 0 | 40 | 100 | . | . | 0.082 |
| Spikevax + Spikevax 14-90 | <5 | 485 | 79 | 43 | 92 | . |
| Spikevax + Spikevax 91-180 | 5 | 594 | 74 | 36 | 89 | . |
| Spikevax + Spikevax 181+ | 17 | 1214 | 60 | 34 | 75 | . |
| Spikevax + Spikevax + Comirnaty 0-13 | <5 | 185 | 81 | -36 | 97 | . |
| Spikevax + Spikevax + Comirnaty 14-60 | <5 | 1709 | 92 | 80 | 97 | . |
| Spikevax + Spikevax + Comirnaty 61+ | 10 | 2076 | 88 | 77 | 94 | . |
| Spikevax + Spikevax + Spikevax 0-13 | <5 | 716 | 84 | 51 | 95 | . |
| Spikevax + Spikevax + Spikevax 14-60 | 8 | 5482 | 96 | 91 | 98 | . |
| Spikevax + Spikevax + Spikevax 61+ | 41 | 5919 | 85 | 78 | 89 | . |
| Vaxzevria 21-83 | 0 | <5 | 100 | . | . | 0.894 |
| Vaxzevria 84+ | 0 | 56 | 100 | . | . | 0.058 |
| Vaxzevria + Vaxzevria 14-90 | 0 | <5 | 100 | . | . | 0.869 |
| Vaxzevria + Vaxzevria 91-180 | <5 | 142 | 37 | -154 | 85 | . |
| Vaxzevria + Vaxzevria 181+ | 12 | 872 | 51 | 12 | 73 | . |
| Vaxzevria + Vaxzevria + Comirnaty 0-13 | <5 | 390 | 78 | 11 | 95 | . |
| Vaxzevria + Vaxzevria + Comirnaty 14-60 | <5 | 2761 | 99 | 91 | 100 | . |
| Vaxzevria + Vaxzevria + Comirnaty 61+ | 12 | 2420 | 86 | 75 | 92 | . |
| Vaxzevria + Vaxzevria + Spikevax 0-13 | <5 | 317 | 89 | 19 | 98 | . |
| Vaxzevria + Vaxzevria + Spikevax 14-60 | <5 | 1531 | 94 | 81 | 98 | . |
| Vaxzevria + Vaxzevria + Spikevax 61+ | 5 | 936 | 87 | 68 | 95 | . |

MLE, maximum likelihood estimate;

LCI/UCI, lower/upper limit of the 95% Wald confidence interval

[1] Likelihood-ratio test

**Supplementary Table 14: Negative control outcome analysis** Ratio comparing the hazard of injury, poisoning and certain other consequences of external causes in vaccinated study subjects with the corresponding hazard in the unvaccinated, adjusted for age, sex, region of residence, residence in a long-term care facility, influenza vaccination in 2019–2020, number of nights hospitalized between 2015 and 2019 and presence of predisposing comorbidities or medical therapies. Estimates marked by an asterisk are statistically significantly different from one.

| | Cases | P-years | MLE | LCI | UCI | p-value[1] | |
|---|---|---|---|---|---|---|---|
| Not vaccinated | 7980 | 243895 | . | . | . | . | |
| Comirnaty 0-20 | 1300 | 40514 | 0.92 | 0.86 | 0.98 | . | * |
| Comirnaty 21-83 | 3704 | 113710 | 0.92 | 0.87 | 0.97 | . | * |
| Comirnaty 84+ | 508 | 9975 | 1.02 | 0.92 | 1.12 | . | |
| Comirnaty + Comirnaty 0-13 | 933 | 26176 | 0.95 | 0.88 | 1.02 | . | |
| Comirnaty + Comirnaty 14-90 | 5129 | 142175 | 0.95 | 0.91 | 1.00 | . | |
| Comirnaty + Comirnaty 91-180 | 5145 | 154744 | 0.95 | 0.90 | 1.01 | . | |
| Comirnaty + Comirnaty 181+ | 1385 | 31053 | 1.07 | 1.00 | 1.16 | . | |
| Comirnaty + Comirnaty + Comirnaty 0-13 | 622 | 20430 | 0.94 | 0.85 | 1.03 | . | |
| Comirnaty + Comirnaty + Comirnaty 14-60 | 2067 | 67674 | 0.97 | 0.90 | 1.04 | . | |
| Comirnaty + Comirnaty + Comirnaty 61+ | 2593 | 72609 | 0.94 | 0.87 | 1.02 | . | |
| Comirnaty + Comirnaty + Spikevax 0-13 | 78 | 2708 | 1.00 | 0.79 | 1.26 | . | |
| Comirnaty + Comirnaty + Spikevax 14-60 | 261 | 8958 | 0.97 | 0.84 | 1.11 | . | |
| Comirnaty + Comirnaty + Spikevax 61+ | 220 | 5434 | 1.14 | 0.98 | 1.33 | . | |
| Spikevax 0-20 | 223 | 4853 | 1.18 | 1.03 | 1.35 | . | * |
| Spikevax 21-83 | 620 | 13710 | 1.14 | 1.04 | 1.24 | . | * |
| Spikevax 84+ | 119 | 1595 | 1.48 | 1.23 | 1.78 | . | * |
| Spikevax + Spikevax 0-13 | 159 | 2997 | 1.27 | 1.08 | 1.49 | . | * |
| Spikevax + Spikevax 14-90 | 861 | 16195 | 1.28 | 1.18 | 1.38 | . | * |
| Spikevax + Spikevax 91-180 | 797 | 17227 | 1.22 | 1.12 | 1.33 | . | * |
| Spikevax + Spikevax 181+ | 172 | 3209 | 1.26 | 1.08 | 1.48 | . | * |
| Spikevax + Spikevax + Comirnaty 0-13 | 26 | 639 | 1.16 | 0.78 | 1.71 | . | |
| Spikevax + Spikevax + Comirnaty 14-60 | 87 | 2077 | 1.22 | 0.98 | 1.52 | . | |
| Spikevax + Spikevax + Comirnaty 61+ | 82 | 2014 | 1.02 | 0.82 | 1.29 | . | |
| Spikevax + Spikevax + Spikevax 0-13 | 83 | 1932 | 1.21 | 0.97 | 1.51 | . | |
| Spikevax + Spikevax + Spikevax 14-60 | 270 | 6380 | 1.21 | 1.06 | 1.39 | . | * |
| Spikevax + Spikevax + Spikevax 61+ | 300 | 5714 | 1.24 | 1.09 | 1.42 | . | * |
| Vaxzevria 0-20 | 57 | 2722 | 0.81 | 0.62 | 1.05 | . | |
| Vaxzevria 21-83 | 227 | 7894 | 1.03 | 0.89 | 1.18 | . | |
| Vaxzevria 84+ | 21 | 469 | 1.48 | 0.96 | 2.27 | . | |
| Vaxzevria + Vaxzevria 0-13 | 36 | 1487 | 0.83 | 0.60 | 1.16 | . | |
| Vaxzevria + Vaxzevria 14-90 | 201 | 8135 | 0.88 | 0.76 | 1.02 | . | |
| Vaxzevria + Vaxzevria 91-180 | 222 | 8915 | 0.99 | 0.86 | 1.14 | . | |
| Vaxzevria + Vaxzevria 181+ | 40 | 1328 | 1.18 | 0.86 | 1.62 | . | |
| Vaxzevria + Vaxzevria + Comirnaty 0-13 | 21 | 901 | 0.98 | 0.64 | 1.51 | . | |
| Vaxzevria + Vaxzevria + Comirnaty 14-60 | 66 | 2985 | 0.94 | 0.73 | 1.20 | . | |
| Vaxzevria + Vaxzevria + Comirnaty 61+ | 52 | 2393 | 0.79 | 0.60 | 1.05 | . | |
| Vaxzevria + Vaxzevria + Spikevax 0-13 | 14 | 463 | 1.22 | 0.72 | 2.07 | . | |
| Vaxzevria + Vaxzevria + Spikevax 14-60 | 42 | 1537 | 1.06 | 0.78 | 1.45 | . | |
| Vaxzevria + Vaxzevria + Spikevax 61+ | 24 | 918 | 0.89 | 0.59 | 1.34 | . | |

MLE, maximum likelihood estimate; LCI/UCI, lower/upper limit of the 95% Wald confidence interval

[1] Likelihood-ratio test

**Supplementary Figure 1: Distribution of SARS-CoV-2 variants by calendar month** A subset of SARS-CoV-2-positive specimens are regularly selected for sequencing to assess the distribution of SARS-CoV-2 variants in Finland. The results are recorded in the National Infectious Diseases Register. The figure shows the relative frequency of each variant between December 27, 2020, and March 31, 2022 in the subpopulation aged 70 years and over. The corresponding number of sequenced specimens as well as the percentage of specimens that were sequenced are given below the x-axis.

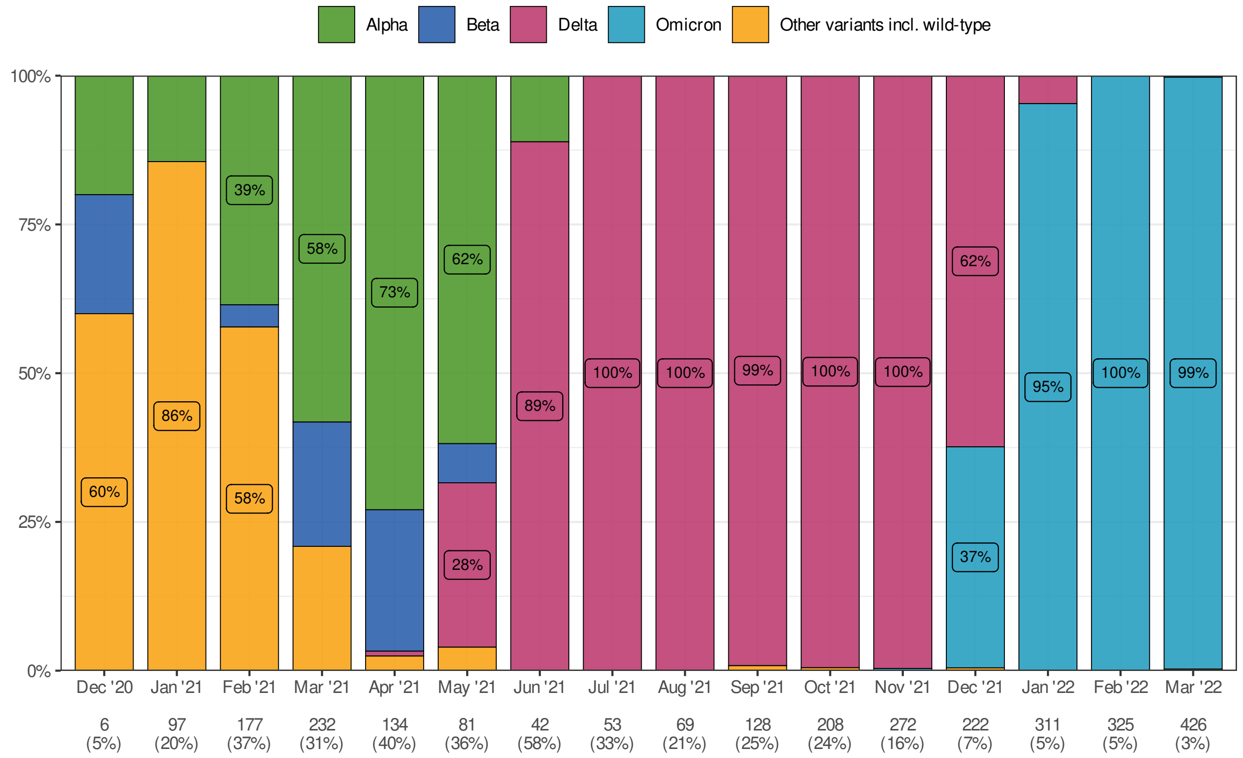

The Omicron subtypes BA.1, BA.2, XJ accounted for 69%, 27%, 4%, respectively, of all Omicron infections.

**Supplementary Figure 2: Distribution of exposure states by calendar month** Relative distribution of person-years in the study of vaccine effectiveness against Covid-19 hospitalization from December 27, 2020, to March 31, 2022.

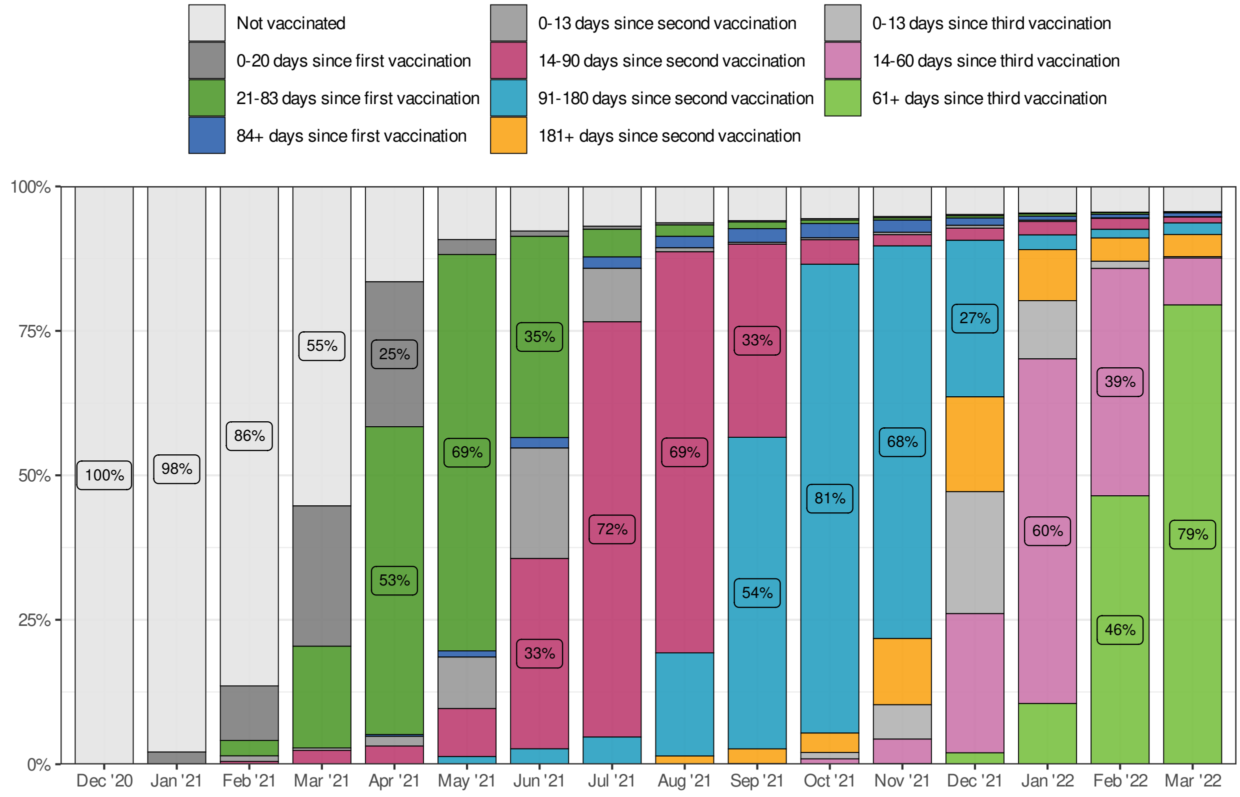

**Supplementary Figure 3: Number of cases by calendar month** Number of Covid-19-related hospital admissions and Covid-19-related ICU admissions in the study, i.e., between December 27, 2020, and March 31, 2022.

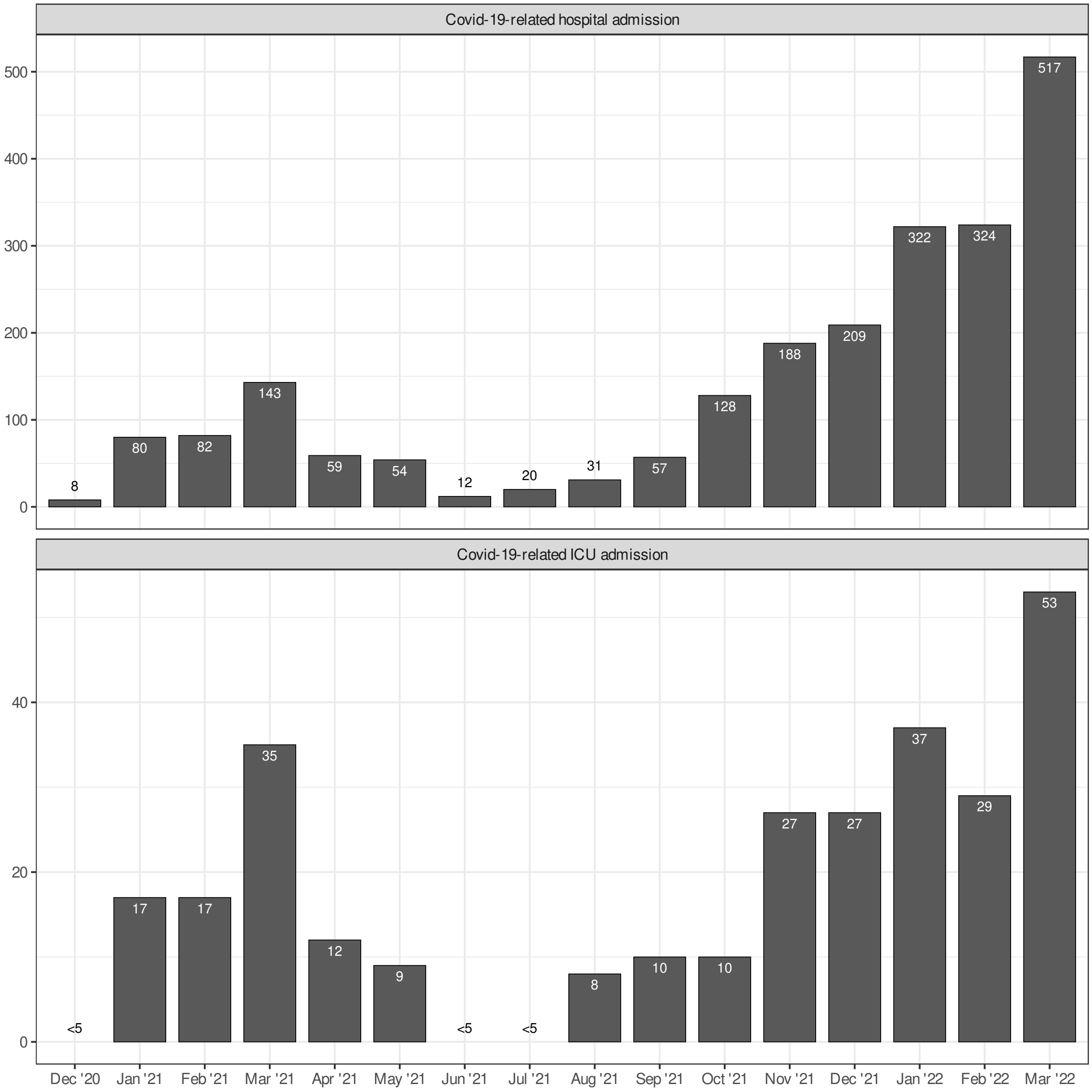

**Supplementary Figure 4: Cumulative hazard of Covid-19-related hospital admission** between December 27, 2020, to March 31, 2022 by calendar time in days and by vaccine, dose and days since last vaccination. For clarity, only the graphs for unvaccinated study subjects and study subjects vaccinated with Comirnaty are shown. The graphs are based on the predicted survival curves adjusted for age, sex, region of residence, residence in a long-term care facility, influenza vaccination in 2019–2020, number of nights hospitalized between 2015 and 2019 and presence of predisposing comorbidities or medical therapies.

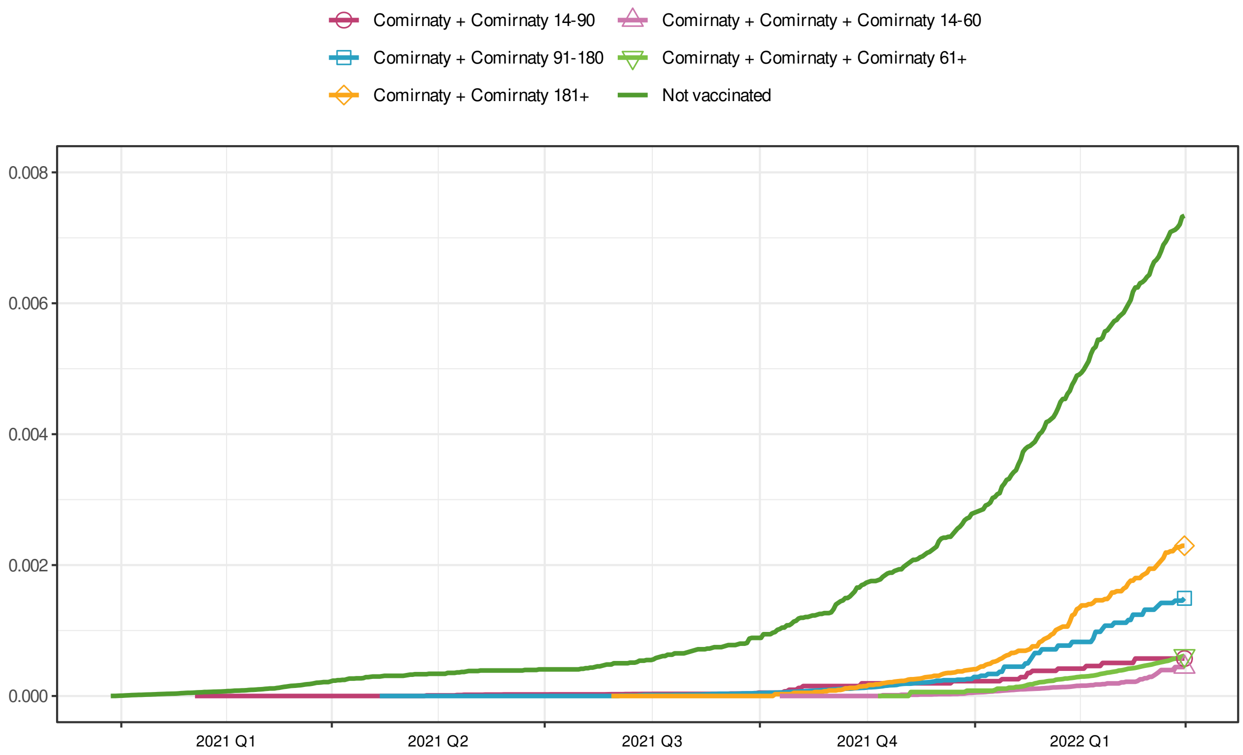

**Supplementary Figure 5: Cumulative hazard of negative control outcome** Cumulative hazard of injury, poisoning and certain other consequences of external causes between December 27, 2020, to March 31, 2022 by calendar time in days and by vaccine, dose and days since last vaccination. For clarity, only the graphs for unvaccinated study subjects and study subjects vaccinated with Comirnaty are shown. The graphs are based on the predicted survival curves adjusted for age, sex, region of residence, residence in a long-term care facility, influenza vaccination in 2019–2020, number of nights hospitalized between 2015 and 2019 and presence of predisposing comorbidities or medical therapies.

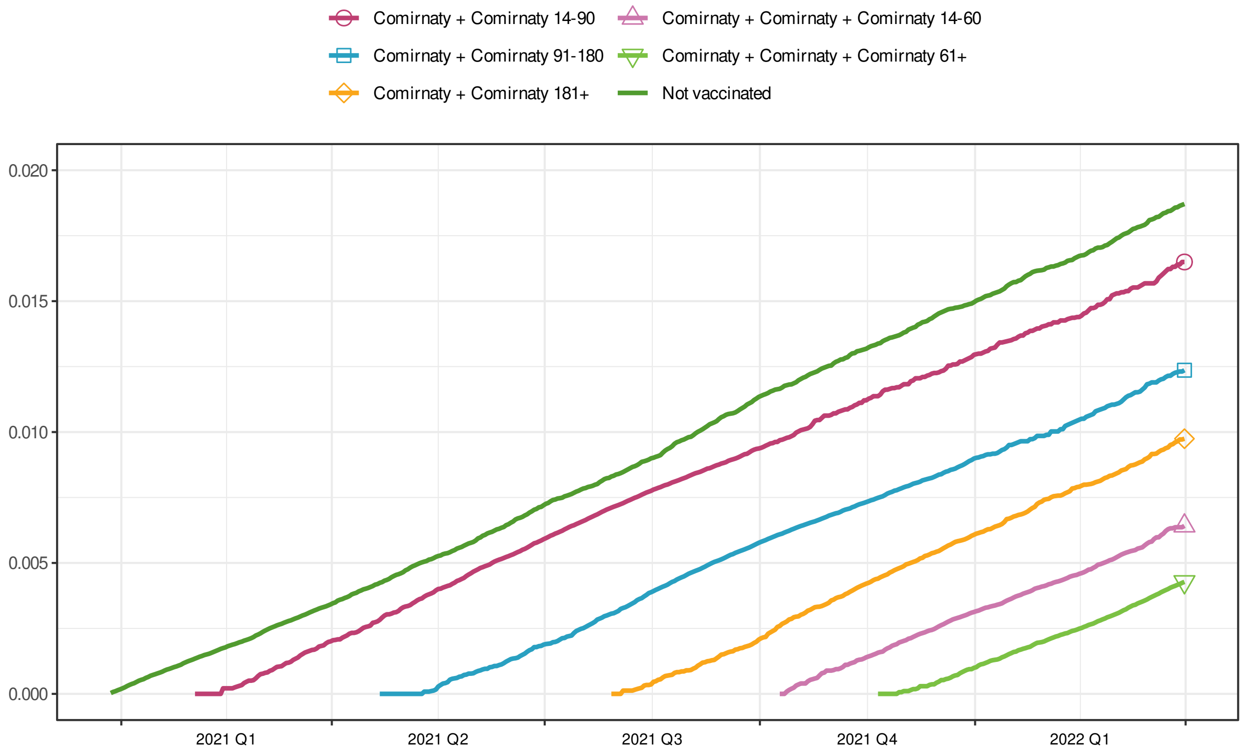
