## Supplementary material for "High vaccine effectiveness against severe Covid-19 in the elderly in Finland before and after the emergence of Omicron": Ethical concern

To whom it may concern:

As director of the department for Health security of the Finnish Institute for Health and Welfare, I certify that:

- I am the competent auhority for assessing whether research requires institutionnal ethical review or if the Finnish communicable diseases law (Tartuntatautilaki 1227/2016)and the law on the duties of the Finnish institute for Health (Laki Terveyden ja hyvinvoinnin laitoksesta 668/2008) and Welfare allows the implementation of the research without seeking further ethical review.

- The research presented by Baum et al in "**High vaccine effectiveness against severe Covid-19 in the elderly in Finland before and after the emergence of Omicron**" did not require further ethical review before implementation as its aim was to monitor vaccine effectiveness of infectious disease (Tartuntatautilaki 1227/2016).

Helsinki, March 3rd 2022
Prof Mika Salminen
